# An Agentic AI Pipeline for an Evidence-Grounded Environmental Health Public Information System: Development and Evaluation

**DOI:** 10.64898/2026.10.26.26366549

**Authors:** Stein H. Ingebretsen, Ram Gouripeddi, Mollie R. Cummins, Julio C. Facelli, Fatemeh Shah-Mohammadi

## Abstract

**Background:** Existing environmental and health information systems typically provide isolated data streams, requiring users to interpret heterogeneous information sources without personalized guidance. Residents and health providers of urban regions with complex environmental conditions, such as the Salt Lake Valley, frequently seek timely, location-specific information about weather, air quality, and their associated health impacts, but may find it difficult to integrate this information in their decision-making, precluding the effective translation of valuable environmental health information into action.

**Objective:** This study aimed to develop and evaluate an agentic artificial intelligence (AI) pipeline that delivers evidence-grounded, neighborhood-scale responses to health-related environmental questions raised lay persons by dynamically orchestrating authoritative weather, neighborhood-scale environmental observations, air quality, and health-evidence resources.

**Methods:** We developed an agentic AI pipeline using LangGraph and OpenAI large language models to interpret natural language queries, select and invoke relevant external tools, retrieve real-time environmental data and evidence-based health guidance, and synthesize responses. The pipeline integrates multiple authoritative resources, including weather forecasts, air-quality observations, geospatial services, and a retrieval-augmented generation (RAG) module built from asthma-related clinical guidance. The system incorporates fallback mechanisms to improve robustness when external services are unavailable. We evaluated the pipeline using a curated benchmark of representative user questions, assessing tool-needed recognition, correct tool selection, avoidance of irrelevant tools, parameter mapping, analytic pathway selection, and incorporation of tool outputs into the final response.

**Results:** The developed system successfully integrated heterogeneous environmental and health information sources into a unified conversational interface that produces evidence-grounded, neighborhood-scale responses for the Salt Lake Valley. Evaluation demonstrated generally strong tool recognition, parameter mapping, and incorporation of retrieved information across diverse question types, supporting the pipeline’s ability to retrieve and integrate authoritative environmental and health information consumable by lay persons and providing an important step for the T3 and T4 phases of the translational pipeline of exposure health informatics.

**Conclusions:** By combining existing environmental health knowledge with local sensor observations and neighborhood-scale environmental measurements, and authoritative state and national weather and air quality resources, the system delivers more localized and context-aware guidance than approaches that rely on authoritative regional or state-level information. Rather than depending on the internal knowledge of a large language model, the pipeline dynamically retrieves, synthesizes, and attributes evidence from authoritative sources. The modular architecture enables seamless integration of additional environmental data streams, health knowledge sources, and geographic regions, providing a scalable foundation for future translational pipelines, completing the T3 to T4 path for environmental health.

## Introduction

Environmental conditions such as air pollution, wildfire smoke, dust events, and temperature inversions have substantial impacts on respiratory health and daily decision-making. These effects are particularly pronounced in regions with complex terrain, like the Salt Lake Valley, where persistent winter inversions, canyon winds, lake-breeze circulations, and seasonal wildfire smoke produce highly heterogeneous environmental conditions across neighborhoods [1–12]. Small spatial and temporal variations in weather and air quality can substantially alter individual exposure, making localized environmental information essential for residents when deciding whether to exercise outdoors, commute, or take preventive measures for respiratory conditions such as asthma. Although state and federal agencies provide authoritative environmental information through weather forecasts, air-quality dashboards, and monitoring networks [1–4,42,43], these resources typically present separate streams of technical data that users must interpret and cooperatively place within the context of their health conditions themselves and often report conditions at regional or county scales rather than at the neighborhood level [53]. This lack of integration and personalization with state-of-the-art environmental health knowledge precludes the translation of primary environmental health research into population-level actionable interventions and behavioral changes, a critical component of the T3 and T4 translational science pathway.

Recent advances in large language models (LLMs) have enabled conversational systems capable of answering complex natural language questions and synthesizing information from multiple sources. Clinical applications have demonstrated the potential of LLMs for knowledge-intensive biomedical tasks, including information extraction, retrieval-augmented generation, clinical decision support, outcome extraction, semantic alignment, and differential diagnosis [17,25–35]. However, conventional LLM-based chatbots rely heavily on parametric knowledge learned during training, which may be outdated, difficult to verify, or insufficient to answer questions requiring current environmental observations. Consequently, responses generated solely from model knowledge may lack provenance, transparency, and the ability to incorporate rapidly changing environmental conditions and emerging environmental health knowledge. To address these limitations, recent research has shifted toward agentic artificial intelligence (AI), in which language models dynamically plan, select, and orchestrate external tools during reasoning rather than relying exclusively on internal model knowledge. Frameworks such as ReAct, Toolformer, TALM, and LangGraph have demonstrated that integrating external tools improves reasoning, factual grounding, and adaptability across diverse tasks [13–16]. More recently, agentic AI has emerged as a promising paradigm for healthcare applications because it enables language models to retrieve authoritative evidence, execute specialized tools, and synthesize information from heterogeneous sources while maintaining traceability of the reasoning process [18–24].

Despite these advances, to our knowledge, there is no system that dynamically integrates authoritative weather forecasts, neighborhood-scale environmental observations, air-quality measurements, and evidence-based environmental health guidance into a unified conversational framework for environmental health question answering by lay persons. This gap is particularly evident in high-altitude mountain areas such as the Salt Lake Valley, where complex terrain, persistent winter inversions, wildfire smoke, dust events, summer heat, and rapidly changing local meteorological conditions create substantial spatial variability in environmental exposures [5–12]. Although authoritative state and federal resources provide high-quality weather forecasts, air-quality information, and environmental observations [1–4,42,43], they are primarily offered as separate dashboards or single-purpose applications. As a result, residents and health professionals must manually interpret and reconcile information from multiple sources to understand their combination effects on health conditions and in the context of their neighborhoods to make informed, and actionable health-related decisions. This precludes the translation of accumulated environmental health research into population-level action.

In this study, we present an agentic AI pipeline that orchestrates multiple authoritative weather, air quality, geospatial, and environmental health evidence resources to answer general and health-related environmental questions. While the pipeline is generic and applicable to any region, we demonstrate its effectiveness for the Salt Lake Valley. The pipeline dynamically selects and invokes the appropriate tools, integrates neighborhood-scale environmental observations with authoritative state and national weather and air-quality products, retrieves evidence-grounded health guidance through retrieval-augmented generation [16,44], and synthesizes evidence-grounded responses with explicit provenance. We evaluate the proposed system using a benchmark of representative user questions, focusing on the agent’s ability to correctly select and invoke tools, extract required parameters, and generate responses grounded in authoritative evidence. Our work demonstrates how agentic AI can bridge heterogeneous environmental information systems to provide transparent, neighborhood-scale environmental health decision support. Unlike conventional environmental dashboards, this system translates heterogeneous environmental and health evidence into patient-oriented information at the point of decision making. The framework provides a model for integrating real-time environmental exposure data with evidence-based health recommendations, creating opportunities for broader deployment of environmental health decision-support systems across translational research networks.

## Methods

The proposed agentic AI system, see Figure 1, is implemented entirely in Python and built on an execution-graph framework, LangChain and LangGraph [16, 45-47]. LangChain provides abstractions and integrations to connect language models with external tools, while LangGraph provides graph-based orchestration to manage state and execution across the agent workflow. The system models user interactions as a minimal state machine that processes user input through tool use and context synthesis. OpenAI GPT-5 model, invoked through an API, serves as the core reasoning agent, interpreting user queries, determining appropriate tool use, and synthesizing retrieved information into the final response.

**Figure 1.**
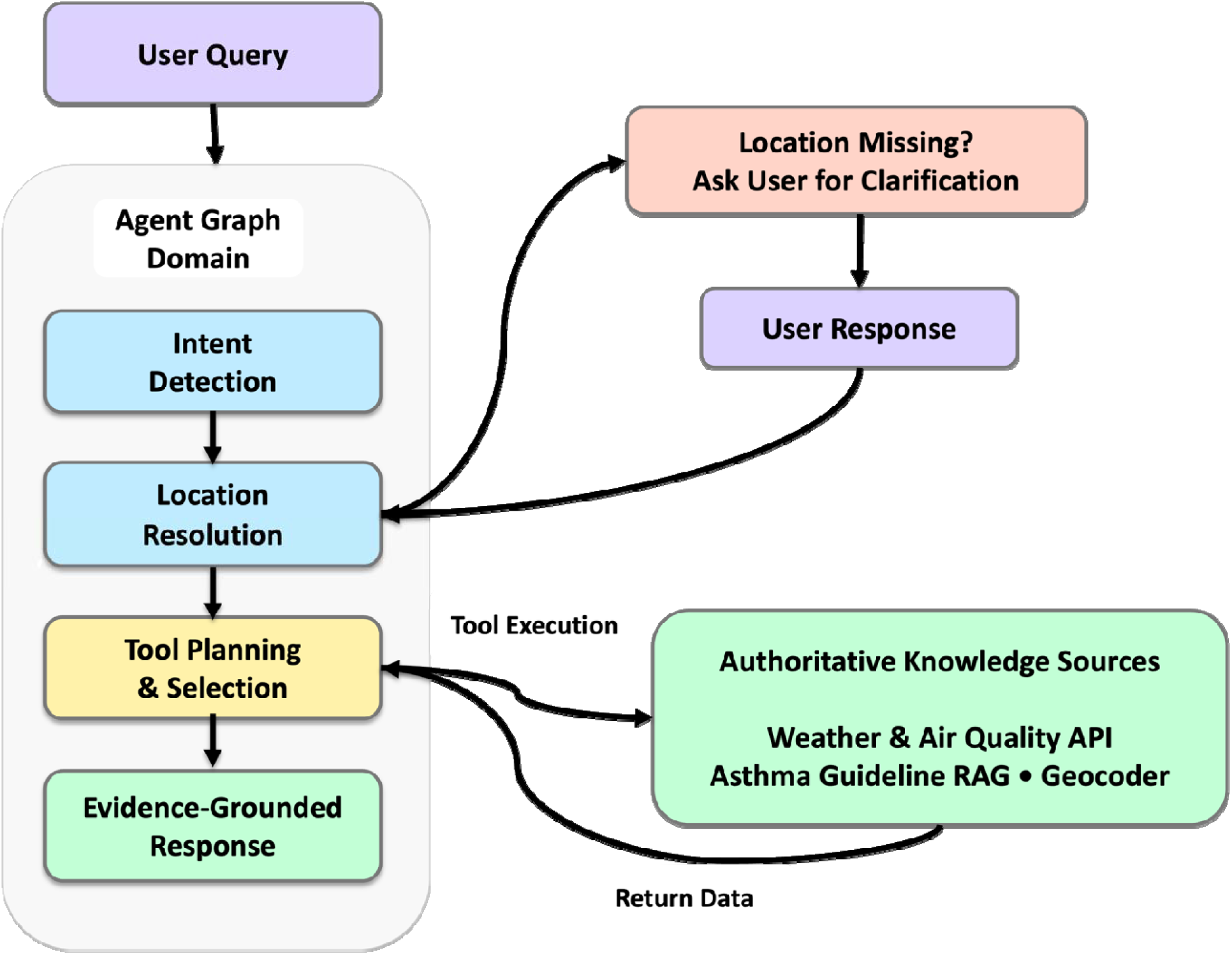
High-level workflow of the agent-based retrieval and response generation system.

### 1 Agent Architecture and Workflow Orchestration

The execution begins by processing the user’s natural language query to identify both the user’s intent and the key entities required to fulfill the request, such as geocoordinate. If sufficient location information is not provided, the agent initiates a clarification dialogue to obtain the missing details before proceeding. Based on the identified intent, the agent dynamically selects and invokes the appropriate external tools, including Retrieval-Augmented Generation (RAG) for evidence-based health information [47,48], weather forecasting services, and local air quality APIs. Table S1 in supplementary materials summarizes the major functional components of the proposed pipeline, including their inputs, outputs, and primary functions. The following subsections describe each component of the pipeline in detail. Once the relevant information has been retrieved, the agent synthesizes the outputs from the selected tools in the context of the user’s query and generates an evidence-grounded response. The final response is formatted in a semi-standardized manner to improve readability and provide users with clear, interpretable environmental and health-related guidance.

#### 1.1 Knowledge Ingestion and Retrieval-Augmented Generation (RAG)

The architecture incorporates a RAG pipeline to support domain-specific knowledge queries. It is designed to maintain structural fidelity across document transformations. Figure 2 provides an overview of the document preparation and indexing pipeline, illustrating the transformation of source documents into structured, retrievable representations for integration into the RAG framework.

**Figure 2.**
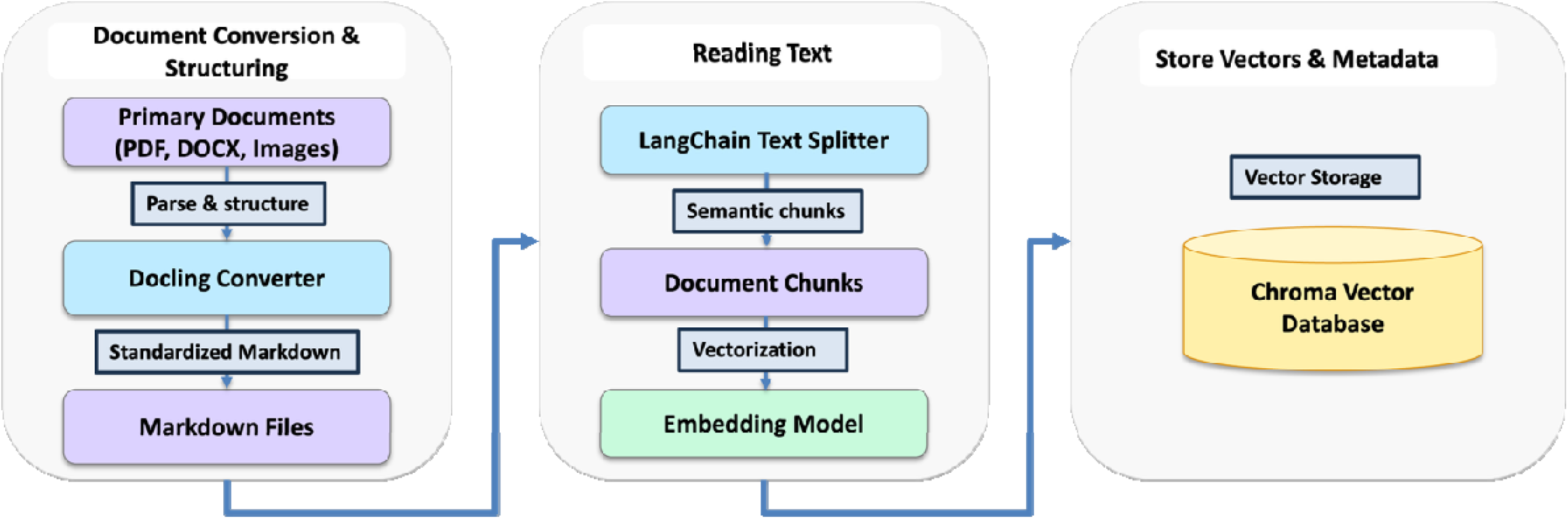
Overview of the document preparation and indexing pipeline.

The document processing pipeline begins by converting unstructured source documents in PDF format into structured Markdown using Docling [49], which preserves textual content, document layout, tables, and other semantic elements, maintaining the original document structure. The resulting Markdown documents are then segmented into semantically meaningful passages using LangChain’s MarkdownTextSplitter, which respects natural document boundaries such as section headings and lists rather than relying on fixed-length text chunks. Each text chunk is subsequently transformed into a dense vector representation using the OpenAI “text-embedding-3-small” embedding model, and the generated embeddings are indexed within a local ChromaDB vector database for efficient cosine-similarity retrieval. Finally, this vector store is integrated into the LangGraph agent as a dedicated retrieval tool, enabling RAG by identifying contextually relevant document segments via cosine-similarity search and providing the retrieved evidence to the agent during response generation. Based on the intent identified in the user’s query, the agent invokes the RAG tool for questions concerning air pollution, ozone, particulate matter, smoke exposure, asthma symptoms, outdoor activity, sensitive populations, or the health implications of weather-related conditions such as extreme heat or cold, humidity, pollen, and thunderstorms. Purely meteorological questions that do not require health-related interpretation or guidance are handled without activating the RAG component. When retrieval is required, the user’s query is converted into an embedding and compared with the stored text-chunk embeddings using cosine similarity. The RAG knowledge base consisted of documents representing complementary clinical and environmental-health evidence. Authoritative sources included the Global Initiative for Asthma (GINA) 2025 “Global Strategy for Asthma Management and Prevention”, the 2020 focused update to the U.S. National Asthma Education and Prevention Program asthma-management guidelines, and the American Lung Association’s “State of the Air 2026” report [60-62]. These were supplemented by peer-reviewed systematic reviews, narrative reviews, meta-analyses, perspectives, and observational and clinical studies examining relationships between asthma and environmental conditions. Collectively, the corpus addressed asthma diagnosis and management, controller and reliever medications, treatment adherence, asthma action plans, symptom and exacerbation management, and modification of outdoor or physical activity. Environmental topics included particulate matter, ozone, nitrogen dioxide, sulfur dioxide, traffic-related and indoor air pollution, smoke, extreme heat and cold, humidity, precipitation, thunderstorms, pollen, climate change, and combined environmental exposures. The literature also provided evidence concerning exposure-related symptoms, lung-function changes, emergency department visits, asthma exacerbations, underlying biological mechanisms, and risks among children, adolescents, adults, and other susceptible populations. The collection was assembled to provide complementary coverage of clinical management, environmental triggers, health effects, vulnerable populations, and practical exposure-reduction strategies relevant to the system’s anticipated weather, air-quality, and asthma-related queries. Given the proof-of-concept scope of this study and its focus on developing and preliminarily evaluating the feasibility of a functional prototype, the RAG corpus was intentionally limited; however, the knowledge base can be expanded to include a larger and more diverse collection of authoritative sources in subsequent stages of development and evaluation.

#### 1.2 External Tool Integration and Resilience Protocols

To integrate real-time observational data, the system incorporates custom API wrappers for air quality and meteorological measurements (Figure 3). Both tools geocode user-supplied locations using the Nominatim geocoding service, accessed through the GeoPy Python client library [54,55]. For weather retrieval, the extracted location is first converted into geographic coordinates (latitude and longitude) using Nominatim’s geocoding service. These coordinates are then used to query the National Oceanic and Atmospheric Administration (NOAA) API [50], which serves as the primary source of localized weather observations and forecast information. To improve system robustness and availability, the weather retrieval pipeline incorporates an automatic fallback mechanism. If the NOAA API request fails due to network timeouts, rate limiting, or other HTTP errors, it is transparently redirected to the Synoptic Data API [51], ensuring continued access to weather observations without requiring additional user interaction. This redundancy improves the agent’s reliability and enables consistent delivery of evidence-grounded responses, even when individual external services are temporarily unavailable.

**Figure 3.**
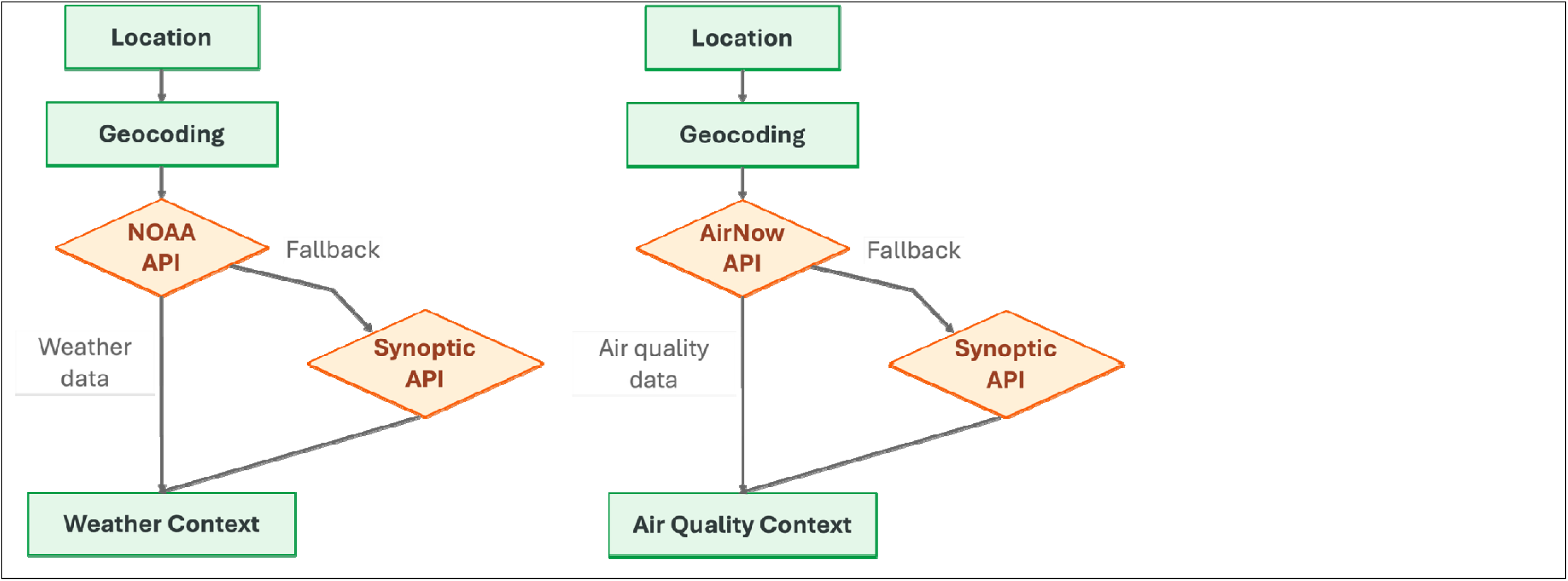
Weather and air quality retrieval workflow.

Similarly, the air quality retrieval pipeline begins by converting the extracted location into geographic coordinates using Nominatim’s geocoding service. The system then queries the AirNow API [52] to real-time ground-level air quality observations, including pollutant-specific measurements and public-health reporting metrics provided by the AirNow platform. Although AQI values are available through AirNow and may be incorporated into responses, the system is not dependent on AQI categories for environmental health reasoning. Instead, the agent combines source pollutant concentrations, meteorological conditions, neighborhood-scale environmental observations, and evidence-based health guidance to generate context-aware recommendations. To ensure reliable data availability, the pipeline incorporates an automatic fallback mechanism that is triggered when the AirNow API is unavailable or when sensor coverage is insufficient for the requested location. In these cases, the system transparently retrieves equivalent observational data from the Synoptic Data API, allowing the agent to continue generating evidence-grounded air quality assessments while minimizing interruptions caused by external service outages or sparse sensor networks.

### 2 Pilot Evaluation Rubric

To evaluate agent execution beyond binary task success, we developed 25 investigator-authored test queries and a domain-specific analytic evaluation rubric. This approach was informed by prior tool-use benchmarks employing expert-authored scenarios and predefined trajectory-level evaluation criteria [56], as well as the Berkeley Function Calling Leaderboard, which uses expert-curated prompts to assess function-calling capabilities across real-world and multi-step agentic settings [57]. However, these general-purpose benchmarks do not address the domain-specific orchestration of weather, air-quality, and RAG tools, or the faithful incorporation of their outputs into environmental-health responses. Therefore, an application-specific test set and evaluation rubric were developed for the proposed pipeline. Each query was evaluated using six binary criteria (0 = not satisfied; 1 = satisfied): (1) recognition of whether external tools or RAG context were required; (2) execution of all scenario-required tools; (3) avoidance of irrelevant tools; (4) accurate extraction and mapping of query entities to tool parameters; (5) routing through the appropriate analytic module or operational pathway, including current-exposure, forecast, fallback, and RAG-only pathways; and (6) faithful incorporation of retrieved API outputs and RAG context into the final response. Criterion scores were summed, yielding a maximum total score of 6 points per query. The sixth criterion provided a limited assessment of final-response fidelity to the retrieved evidence. The team manually reviewed the trajectories of the tools employed for each question and the final responses; all the responses were found to be factually correct to the contents provided in the knowledge-base (rag-based source) and in the context of location and time of the questions. Therefore, the evaluation characterizes the system’s technical and operational performance in delivering contextual responses, rather than establishing its clinical validity. The pilot benchmark comprised 25 representative queries (Q01-Q25) covering diverse real-world requests (e.g., current conditions, multi-day forecasting, atmospheric dynamics, and domain-specific guideline retrieval). The complete set of benchmark questions is provided in the Supplementary Materials to facilitate reproducibility and enable future comparative evaluations of similar agentic AI systems. The agent’s step-by-step execution logs and generated responses were recorded and scored against the 6-point rubric.

## Results

The pilot evaluation included 25 representative user queries spanning current weather conditions, multi-day forecasting, air quality dynamics, and evidence-based health guidance retrieval. The complete list of evaluation questions and the details of the scores per question against the 6-point rubric are provided in the Supplementary Materials Table S2 and S3. A representative execution trace for second user query (Q02), illustrating the agent’s use of AirNow air-quality retrieval and asthma-guideline RAG to generate a final response, is shown in Supplementary Materials Figure S1.

**Table 1.** Descriptive Statistics of Tool-Calling Performance Across the Pilot Benchmark.

| Statistic | Tool-needed recognition | Correct required tool selection | Avoidance of irrelevant tools | Correct parameter mapping | Correct analytic module or guideline pathway | Correct incorporation of tool outputs into final answer |
| --- | --- | --- | --- | --- | --- | --- |
| Mean | 0.96 | 0.80 | 1 | 0.92 | 0.8 | 0.96 |
| Median | 1 | 1 | 1 | 1 | 1 | 1 |
| Mode | 1 | 1 | 1 | 1 | 1 | 1 |
| StDev | 0.20 | 0.408 | 0 | 0.277 | 0.408 | 0.20 |
| SEM | 0.04 | 0.082 | 0 | 0.055 | 0.082 | 0.04 |
| 95% Margin of Error | 0.083 | 0.169 | N/A | 0.144 | 0.169 | 0.083 |

*\*Note: 95% Margin of Error (MOE) is undefined/zero due to zero variance across all observations.*

**Table 2.**
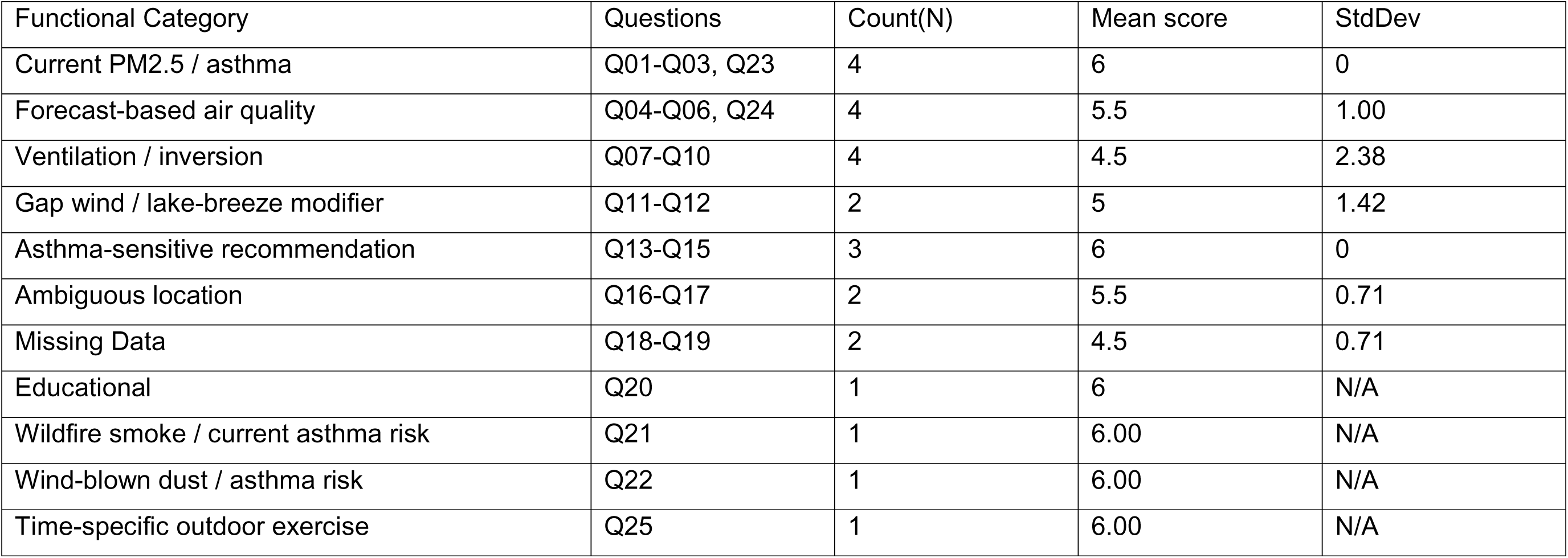
Tool-Calling Performance by Functional Category Across the Pilot Benchmark.

Across the 25 benchmark queries, the agentic framework achieved an aggregate score of 136 out of 150 possible rubric points, representing an overall compliance rate of 90.7%. The agent demonstrated strong performance across the tool-calling workflow, correctly recognizing when external tools were required for 96% of queries and successfully incorporating retrieved tool outputs into the final response for 96% of cases. The system consistently avoided invoking unnecessary tools, achieving perfect performance (100%) on irrelevant tool avoidance across all benchmark questions. Correct parameter mapping, including extraction of location-related entities required for API execution, was achieved in 92% of evaluations (mean=0.92). Lower performance was observed for correct selection of the required tool and for selecting the appropriate analytic module or guideline pathway, both of which achieved an average accuracy of 80%. These errors primarily occurred in a small number of questions where the agent either selected an inappropriate retrieval pathway or failed to invoke the expected domain-specific tool despite correctly recognizing that tool use was necessary.

Across all evaluation dimensions, the median and modal performance corresponded to perfect scores, indicating that most benchmark questions were handled successfully.

Table 2 summarizes the performance of the proposed agentic AI pipeline across 11 functional categories represented in the 25-query pilot benchmark. The categories reflect common environmental health information needs, including current and forecast air quality, meteorological conditions, asthma-sensitive recommendations, location ambiguity, missing data, wildfire smoke, wind-blown dust, and time-specific outdoor exercise. The agent achieved perfect performance (mean score=6.00; SD=0) for Current PM_2.5_/Asthma and Asthma-Sensitive Recommendation queries. Educational, Wildfire Smoke/Current Asthma Risk, Wind-Blown Dust/Asthma Risk, and Time-Specific Outdoor Exercise queries also each received a perfect score of 6.00; These results indicate consistent tool selection, parameter extraction, and evidence-grounded response generation for these question types. Forecast-Based Air Quality queries demonstrated strong performance (mean=5.50; SD=1.00), although occasional errors occurred in selecting the appropriate forecasting tool or analytic pathway. Lower performance was observed for Ventilation/Inversion and Missing Data questions, both of which achieved a mean score of 4.50. The relatively high standard deviation for the Ventilation/Inversion category (SD=2.38) indicates greater variability in performance, reflecting the complexity of queries that require integration of multiple meteorological variables and specialized reasoning pathways. Missing Data scenarios similarly highlighted opportunities to improve fallback strategies and response generation when external information is incomplete. Gap Wind/Lake-Breeze Modifier questions achieved a mean score of 5.00 (SD=1.42), while Ambiguous Location queries achieved a mean score of 5.50 (SD=0.71). These findings suggest that the clarification dialogue and location-resolution components were generally effective, with occasional failures in parameter mapping or downstream tool selection. Overall, the category-level analysis indicates that the pipeline performs reliably across a broad range of environmental health question types while identifying complex meteorological reasoning and incomplete-data scenarios as the primary areas for future improvement. Following human expert review of the pipeline trajectories and final answers, all 25 responses were judged factually accurate within the available tools and the supplied spatiotemporal context [58].

## Discussion

This work addresses a translational gap between environmental health evidence and community-level decision making. The system operationalizes environmental observations and evidence-based asthma guidance within an actionable conversational interface, representing a T3 implementation approach. By delivering evidence-grounded recommendations to residents and healthcare stakeholders in real-world settings, the framework also establishes infrastructure for future T4 evaluation studies of adoption, behavior change, and population-level health outcomes. Unlike AQI alone, which was designed primarily as a public communication tool, the proposed framework combines environmental observations with disease-specific guidance and contextual risk interpretation to support patient-centered decisions.

An important consideration is the role of AQI in environmental health risk communication. Traditional AQI frameworks communicate population-level pollution alerts and are not intended to represent personalized health risk. AQI reporting typically emphasizes the pollutant contributing to the highest index value at a given time, which can obscure concurrent exposure to multiple pollutants and their cumulative effects [53]. In addition, people with asthma and other chronic respiratory diseases may experience adverse effects at pollutant concentrations considered acceptable within regulatory frameworks [59]. Although the proposed system currently incorporates AQI-reporting data sources, its modular architecture can extend beyond AQI-based communication by integrating pollutant observations, meteorological context, local environmental conditions, and evidence-based asthma guidance. This capability is an important step toward individualized environmental health decision support.

One limitation emerged during forecast-oriented air quality queries, particularly those requesting next-day or future air quality conditions. In these scenarios, the agent correctly recognized the need for environmental information and successfully invoked the appropriate air quality retrieval tool. However, the integrated AirNow API provides real-time observational measurements rather than predictive air quality forecasts. Because the orchestration logic did not distinguish between observational and forecast data, the retrieved real-time air quality values were incorporated directly into responses addressing future conditions. As a result, the system incorrectly interpreted current observations as future predictions. This finding highlights an important temporal reasoning limitation within the current agent architecture, where successful tool selection alone is insufficient to ensure that retrieved information matches the temporal intent of the user’s query. We addressed this issue by incorporating temporal consistency checks into the orchestration workflow and integrating forecast-specific air quality services and predicted air quality data when responding to predictive environmental health questions. Consequently, environmental risk characterization remains partially dependent on conventional regulatory reporting frameworks. Future work will evaluate incorporation of multipollutant exposure indices, cumulative exposure models, environmental mixtures approaches, and personalized susceptibility profiles to provide more individualized environmental health assessments [53]. Since this work is a proof-of-concept study, the evaluation was limited to 25 domain-specific queries, in contrast to larger general-purpose benchmarks [56]. Accordingly, the findings should be interpreted as preliminary evidence of technical feasibility rather than a comprehensive or definitive assessment of agent performance. Future evaluations should include a substantially larger, independently developed question set; repeated executions; external expert review; and testing with real users.

## Conclusions

In this study, we developed an agentic AI system that integrates real-time weather and air quality data with evidence-based health guidance for asthma to provide localized, context-aware responses to natural language queries. Using LangChain and LangGraph, the system dynamically interprets user intent, resolves geographic locations, selects appropriate tools, and synthesizes outputs from authoritative environmental data sources and a retrieval-augmented knowledge base. This architecture demonstrates the feasibility of combining graph-based orchestration, real-time environmental monitoring, and domain-specific knowledge retrieval within a unified conversational system, while providing a foundation for future evaluation and deployment of reliable environmental health decision-support tools. The modular architecture enables seamless integration of additional environmental data streams, health knowledge sources, adaptation to other diseases and geographic regions, providing a scalable foundation for future environmental health decision-support applications. The modular architecture enables seamless integration of additional environmental data streams, health knowledge sources, and geographic regions, providing a scalable foundation for future translational pipelines completing the T1 to T4 path for environmental health.

## Ethics Approval

No protected health information was collected or used for this work.

## Data Availability Statement

The data underlying this article are available at: https://zenodo.org/records/22286093

## Supplementary Materials

**Table S1.**
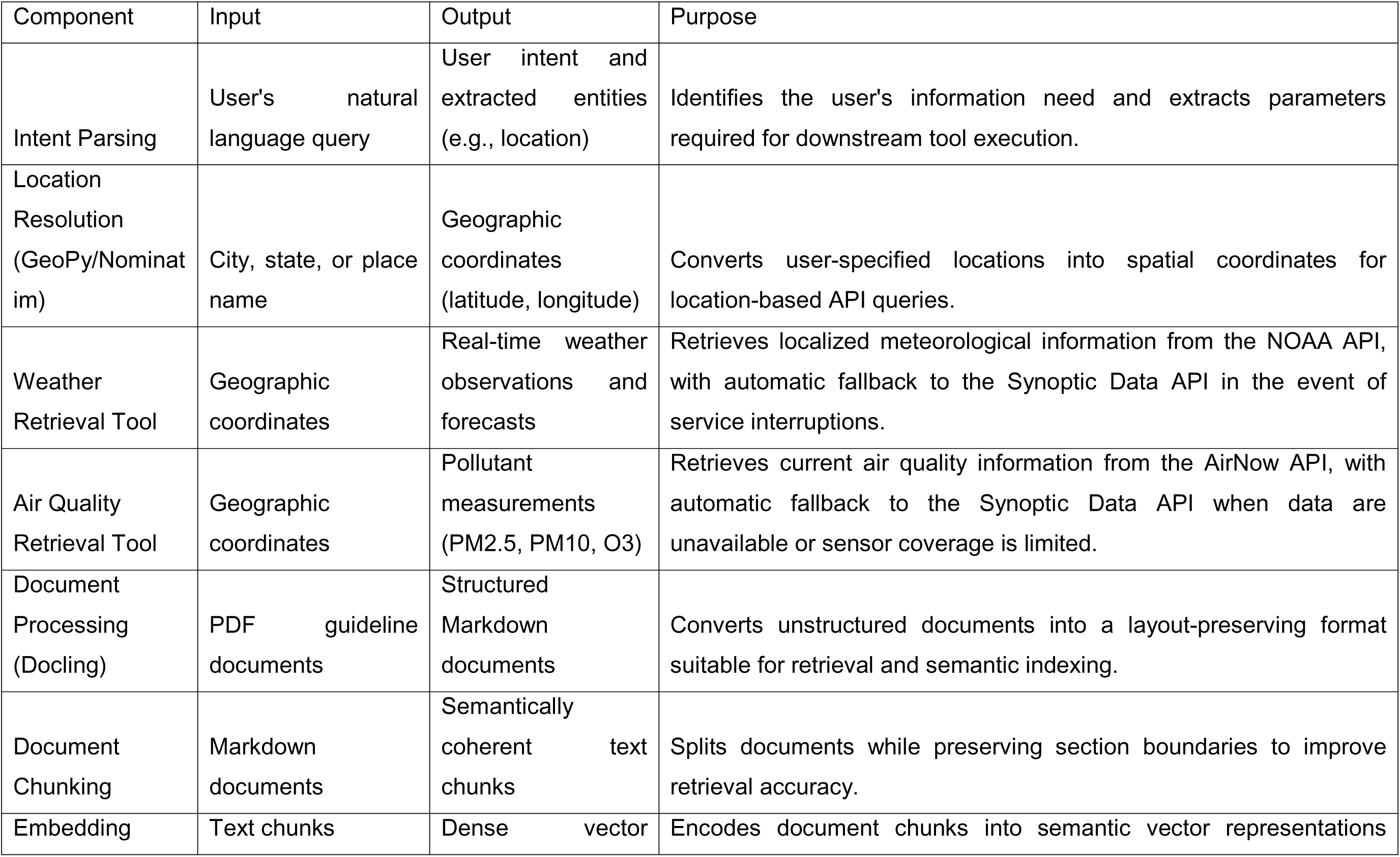
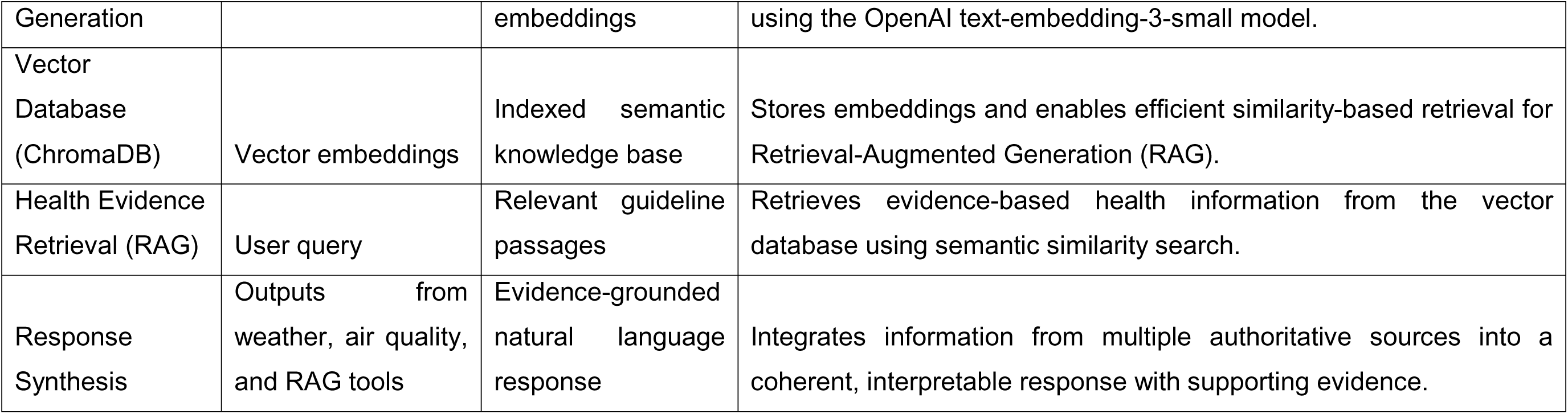
Functional components of the agentic AI pipeline.

**Table S2:**
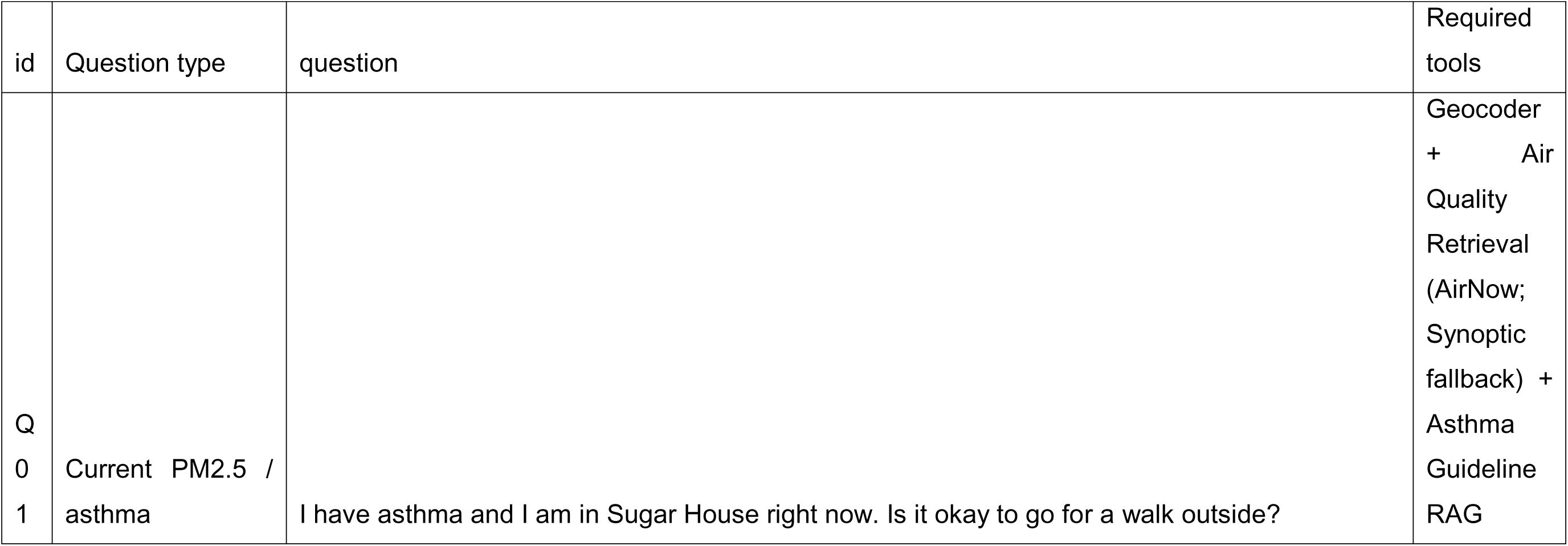
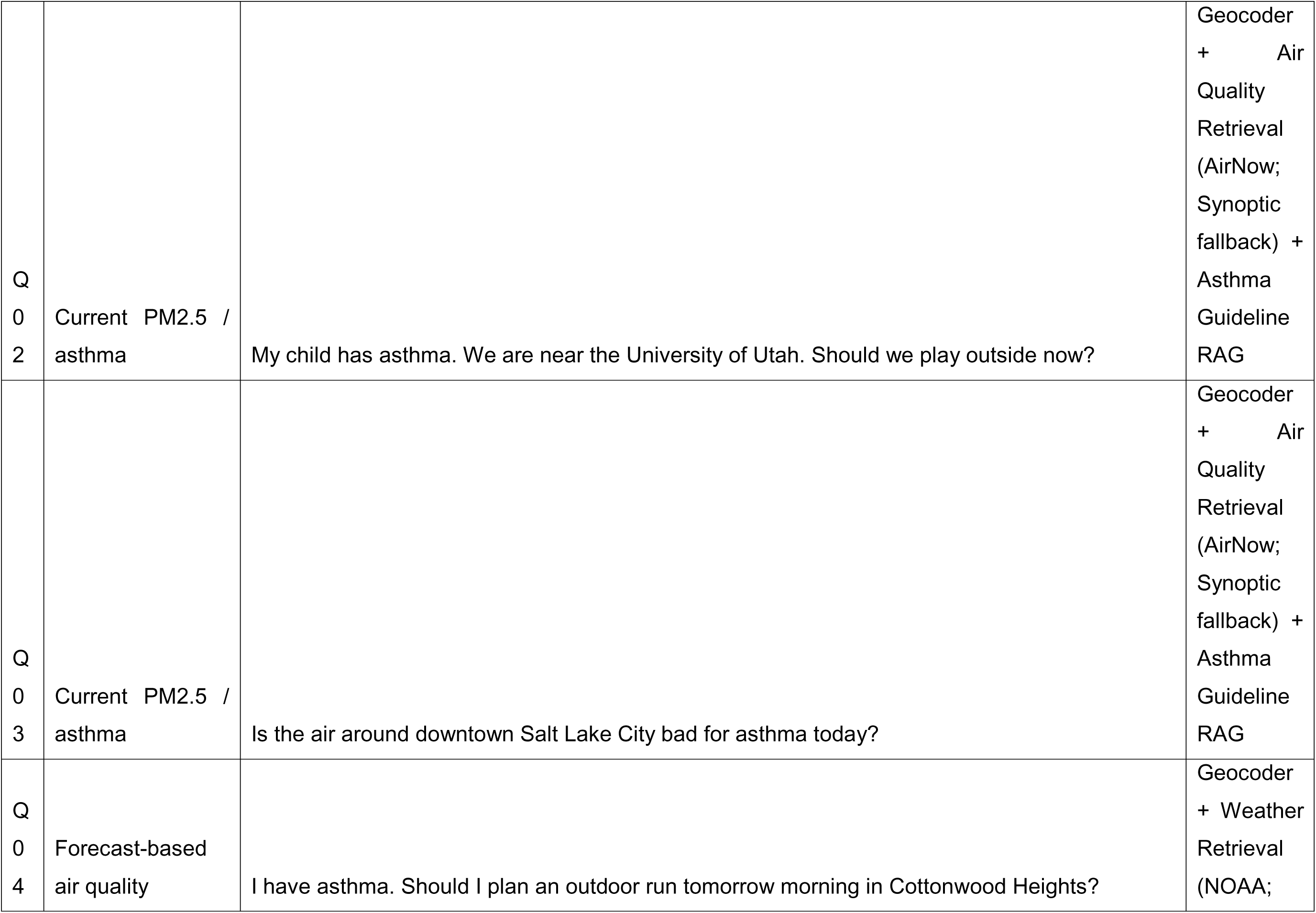
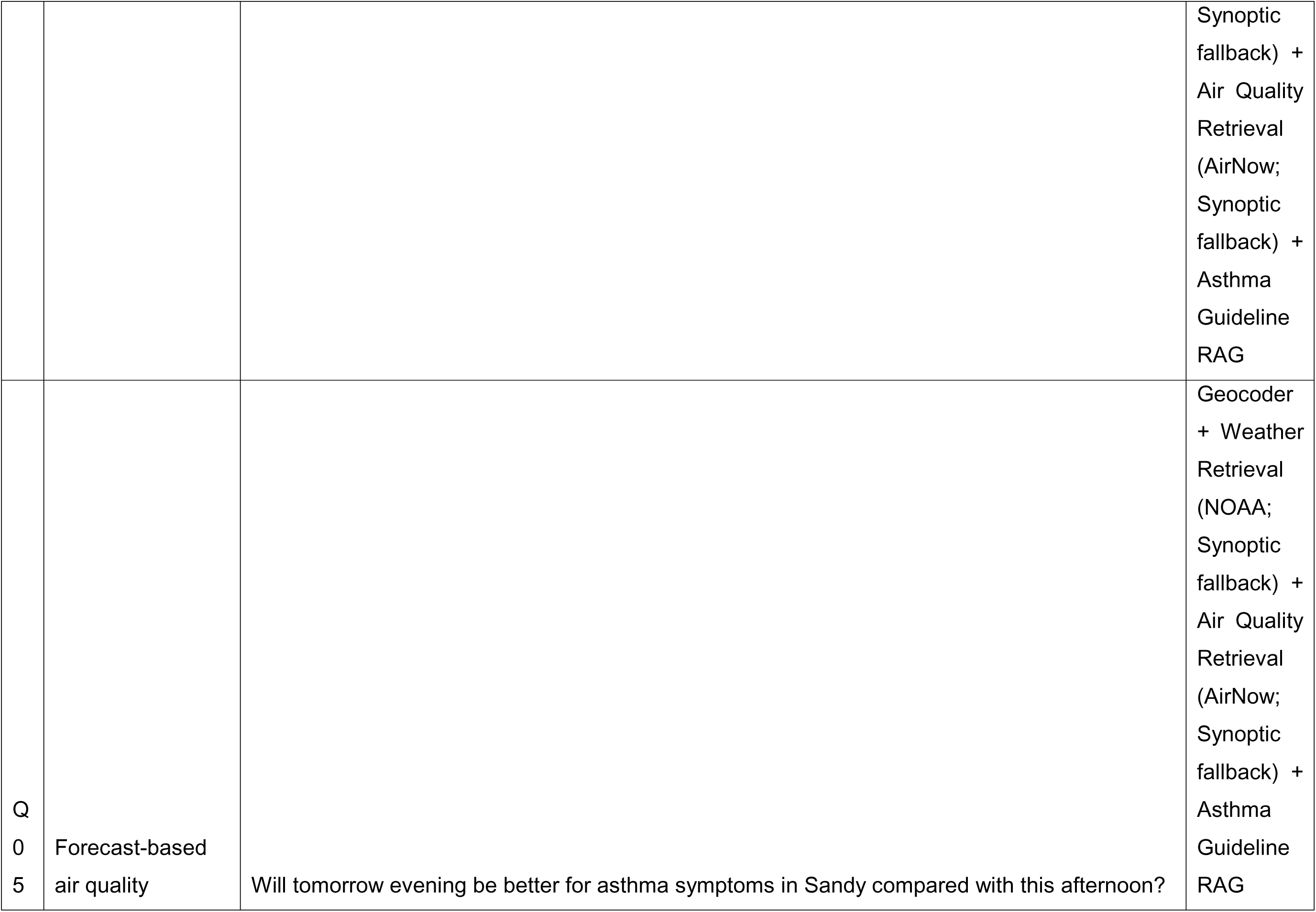
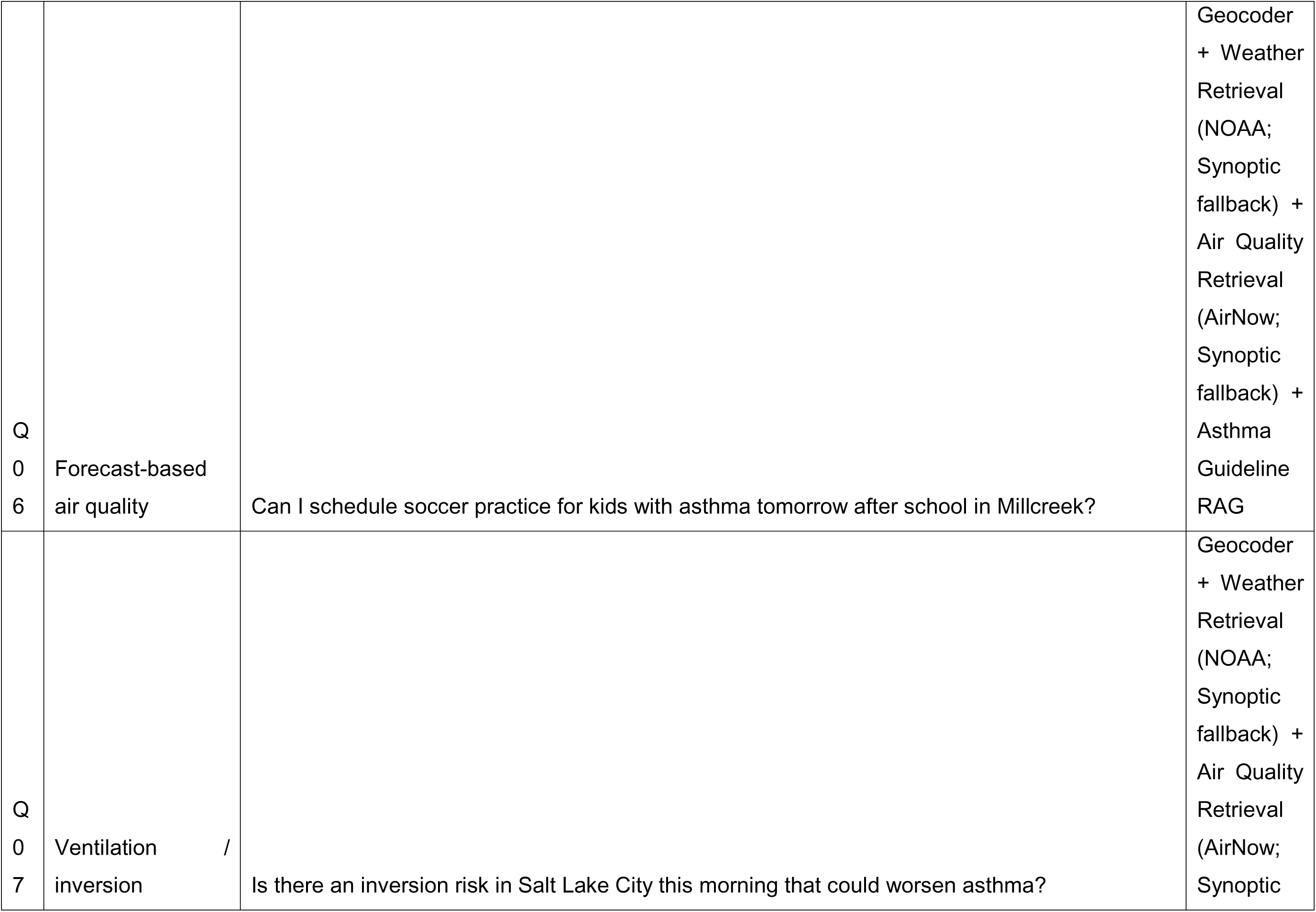
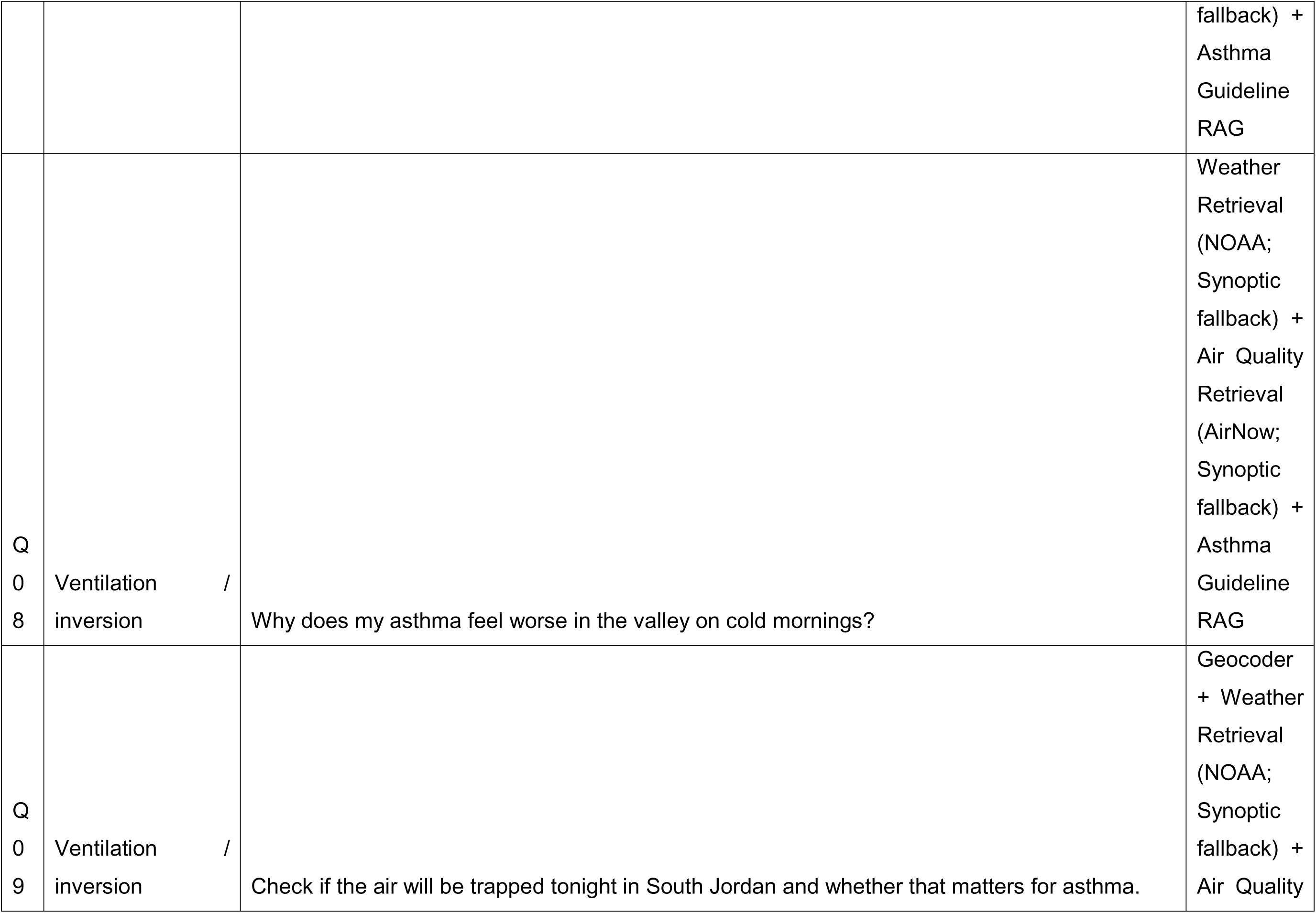
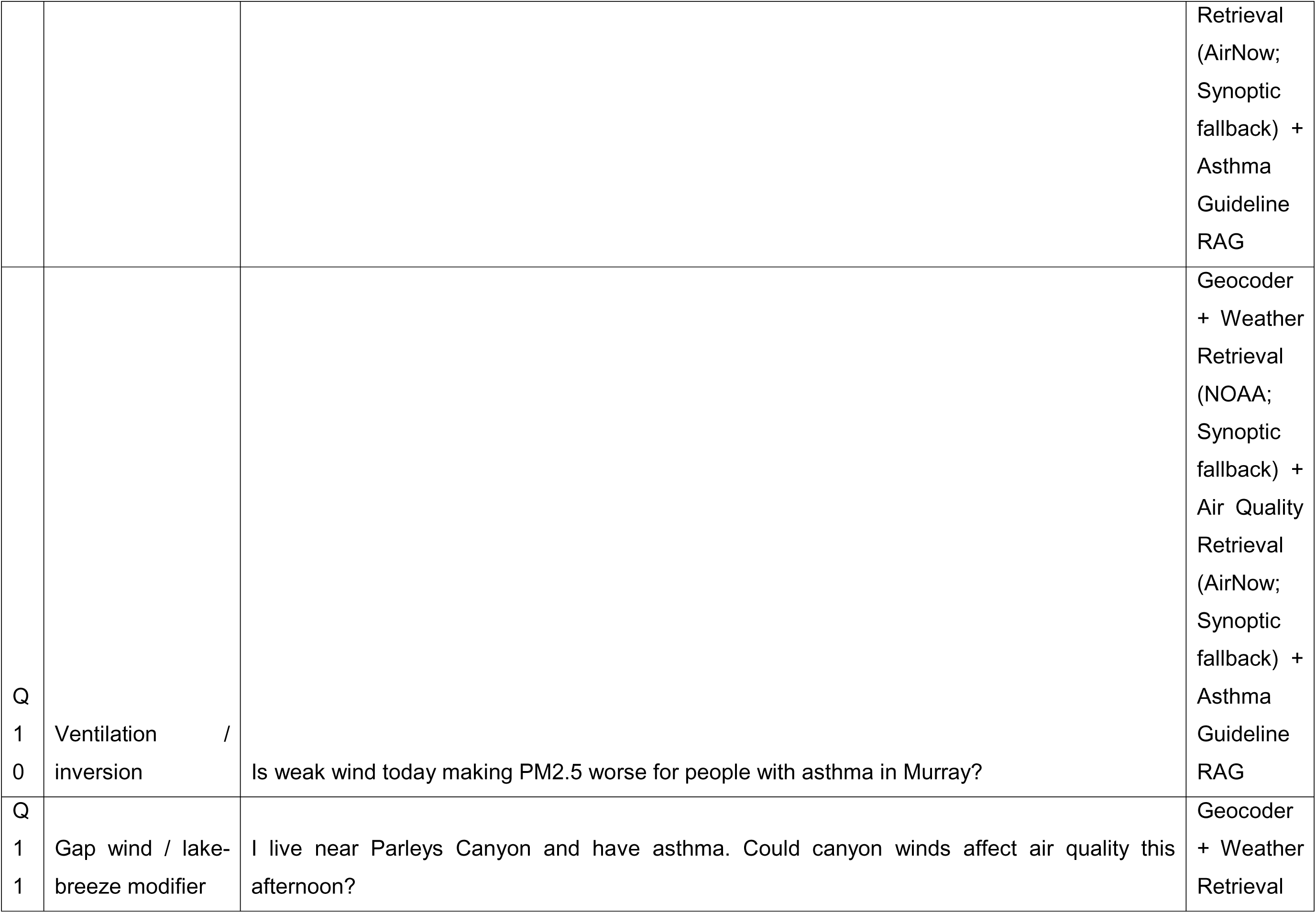
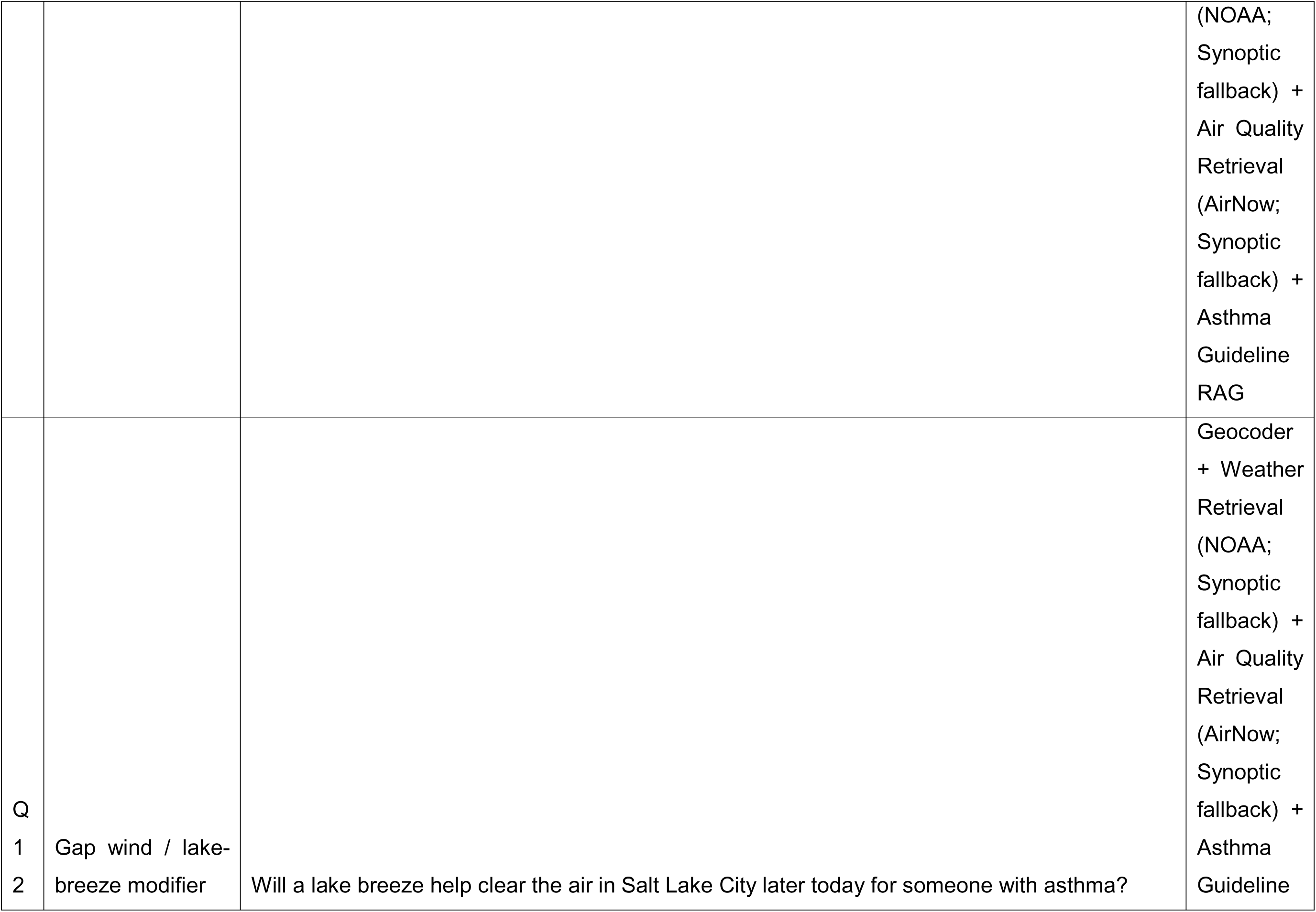
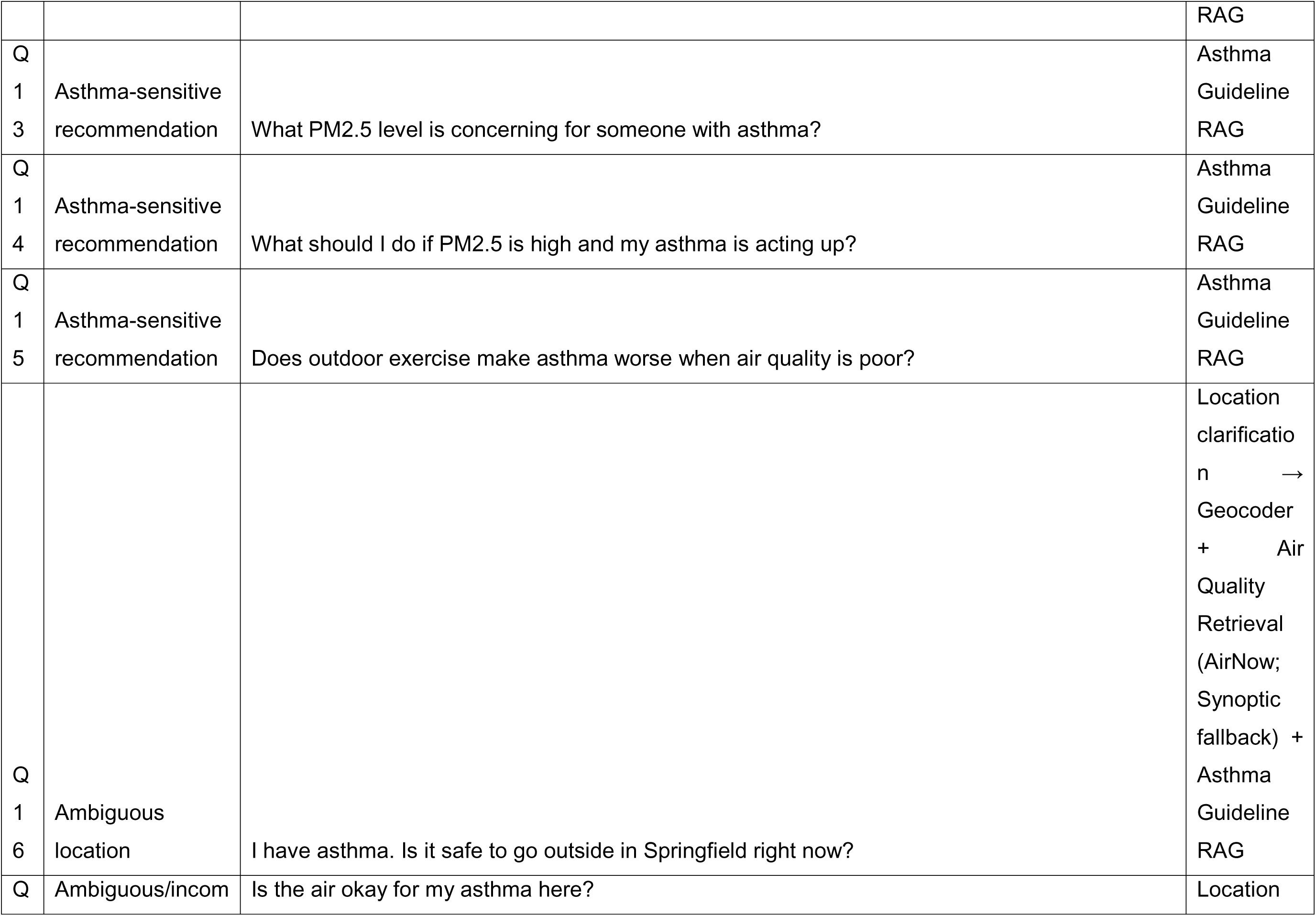
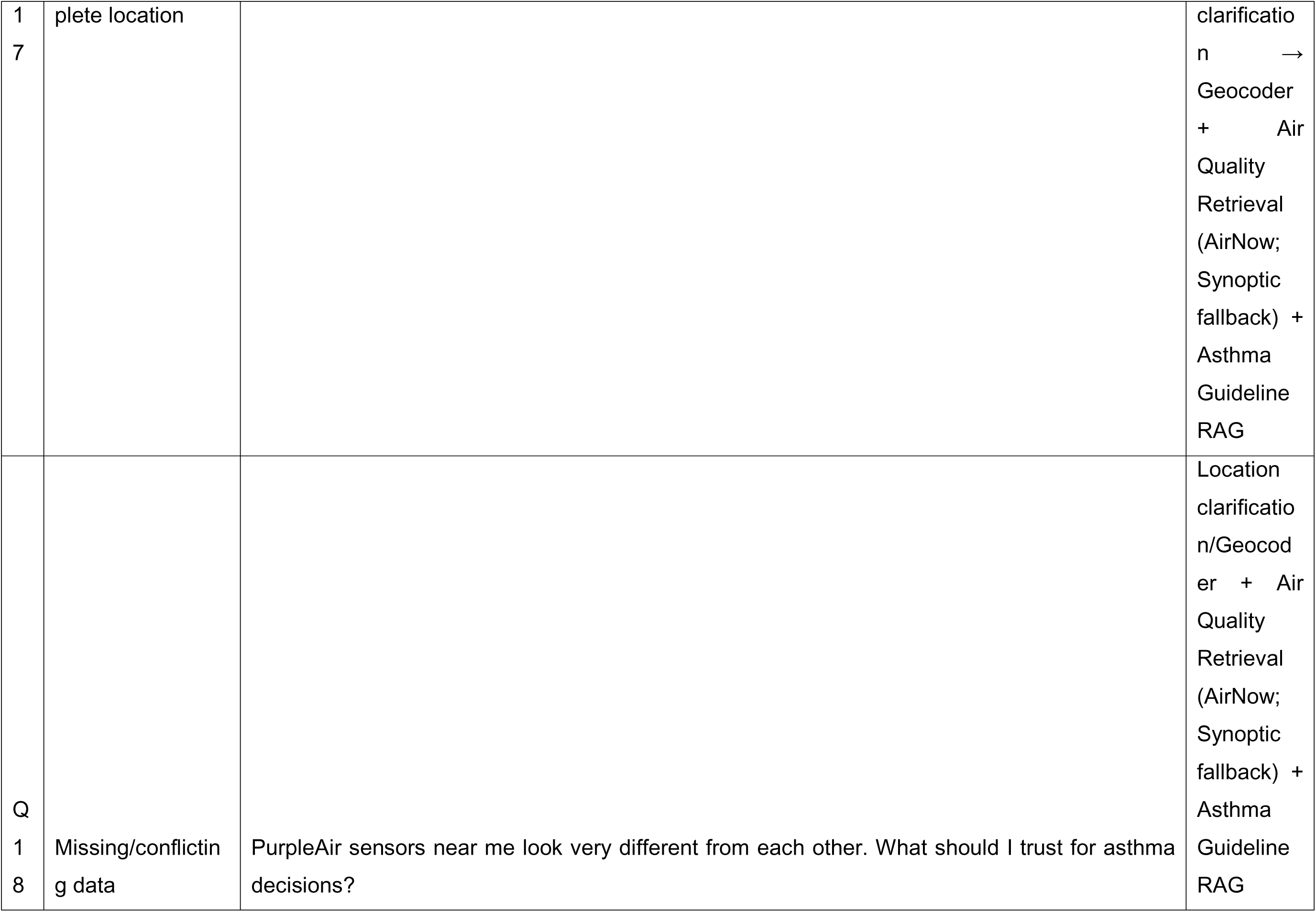
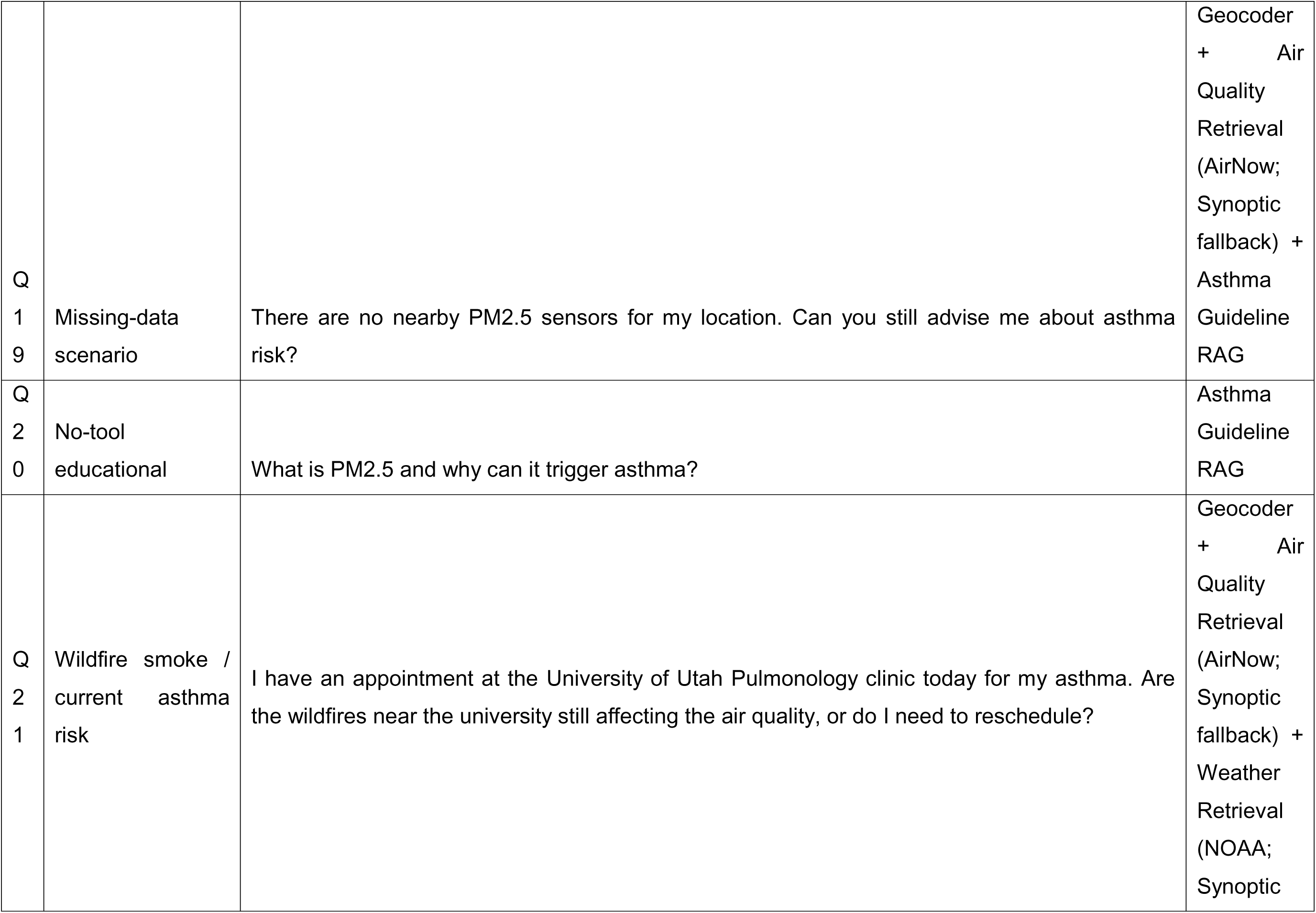
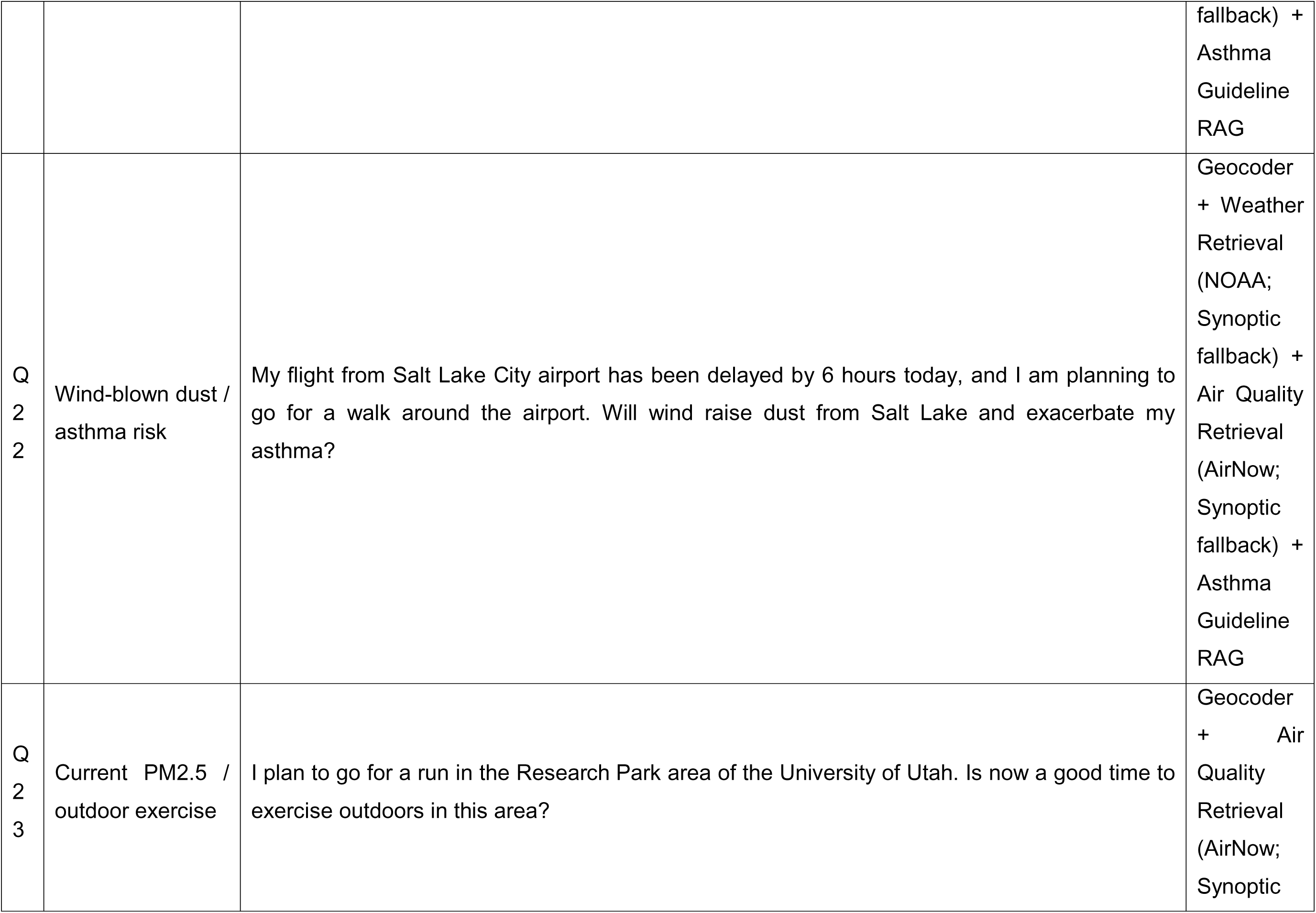
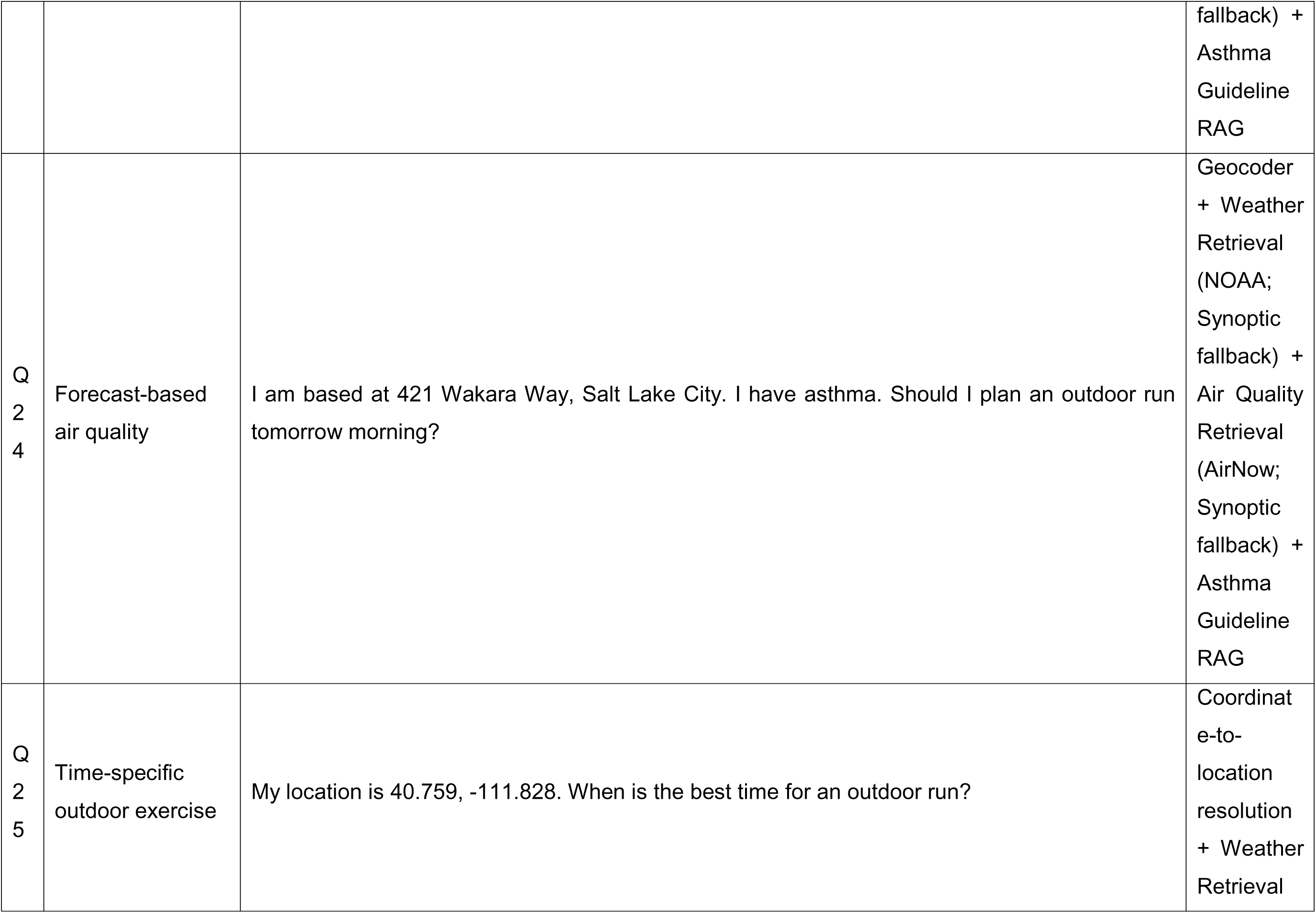
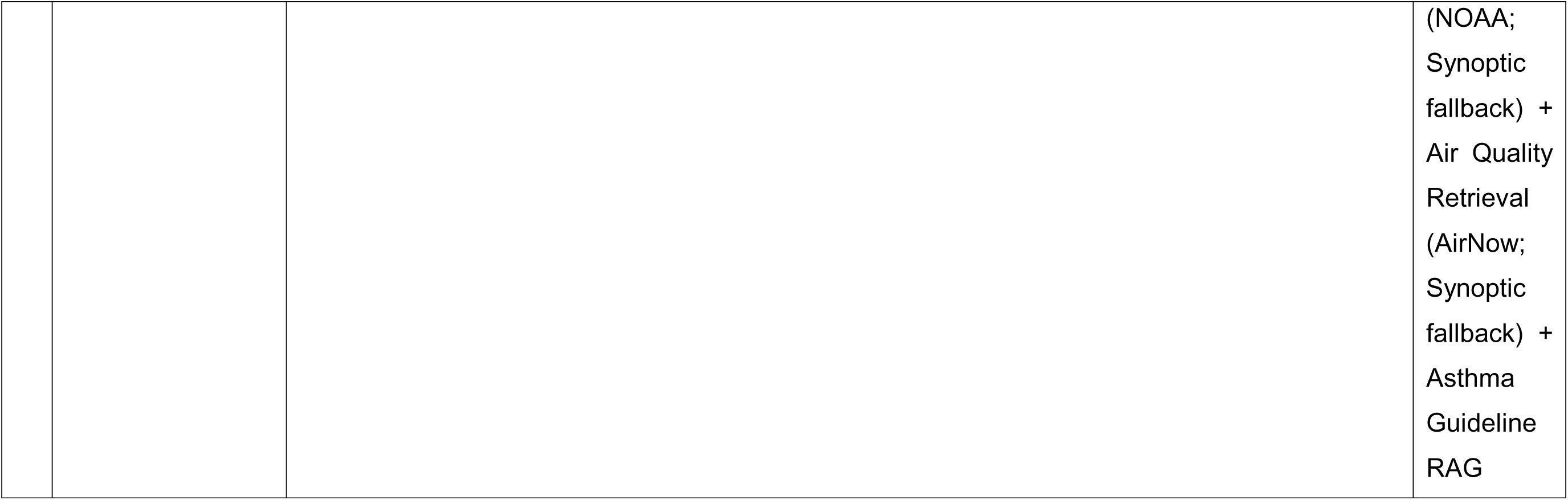
Benchmark question set and required tools.

**Table S3:** Functional components of the agentic AI pipeline.

| Que<br>stion | Tool-needed<br>recognition | Correct<br>required tool<br>selection | Avoidance of<br>irrelevant tools | Correct<br>parameter<br>mapping | Correct analytic module<br>or guideline pathway | Correct incorporation of tool<br>outputs into final answer | To<br>tal |
| --- | --- | --- | --- | --- | --- | --- | --- |
| Q01 | 1 | 1 | 1 | 1 | 1 | 1 | 6 |
| Q02 | 1 | 1 | 1 | 1 | 1 | 1 | 6 |
| Q03 | 1 | 1 | 1 | 1 | 1 | 1 | 6 |
| Q04 | 1 | 1 | 1 | 1 | 1 | 1 | 6 |
| Q05 | 1 | 1 | 1 | 1 | 1 | 1 | 6 |
| Q06 | 1 | 0 | 1 | 1 | 0 | 1 | 4 |
| Q07 | 1 | 0 | 1 | 1 | 1 | 1 | 5 |
| Q08 | 0 | 0 | 1 | 0 | 0 | 0 | 1 |
| Q09 | 1 | 1 | 1 | 1 | 1 | 1 | 6 |
| Q10 | 1 | 1 | 1 | 1 | 1 | 1 | 6 |
| Q11 | 1 | 0 | 1 | 1 | 0 | 1 | 4 |
| Q12 | 1 | 1 | 1 | 1 | 1 | 1 | 6 |
| Q13 | 1 | 1 | 1 | 1 | 1 | 1 | 6 |
| Q14 | 1 | 1 | 1 | 1 | 1 | 1 | 6 |
| Q15 | 1 | 1 | 1 | 1 | 1 | 1 | 6 |
| Q16 | 1 | 1 | 1 | 0 | 1 | 1 | 5 |
| Q17 | 1 | 1 | 1 | 1 | 1 | 1 | 6 |
| Q18 | 1 | 1 | 1 | 1 | 0 | 1 | 5 |
| Q19 | 1 | 0 | 1 | 1 | 0 | 1 | 4 |
| Q20 | 1 | 1 | 1 | 1 | 1 | 1 | 6 |
| Q21 | 1 | 1 | 1 | 1 | 1 | 1 | 6 |
| Q22 | 1 | 1 | 1 | 1 | 1 | 1 | 6 |
| Q23 | 1 | 1 | 1 | 1 | 1 | 1 | 6 |
| Q24 | 1 | 1 | 1 | 1 | 1 | 1 | 6 |
| Q25 | 1 | 1 | 1 | 1 | 1 | 1 | 6 |

**Figure S1:**
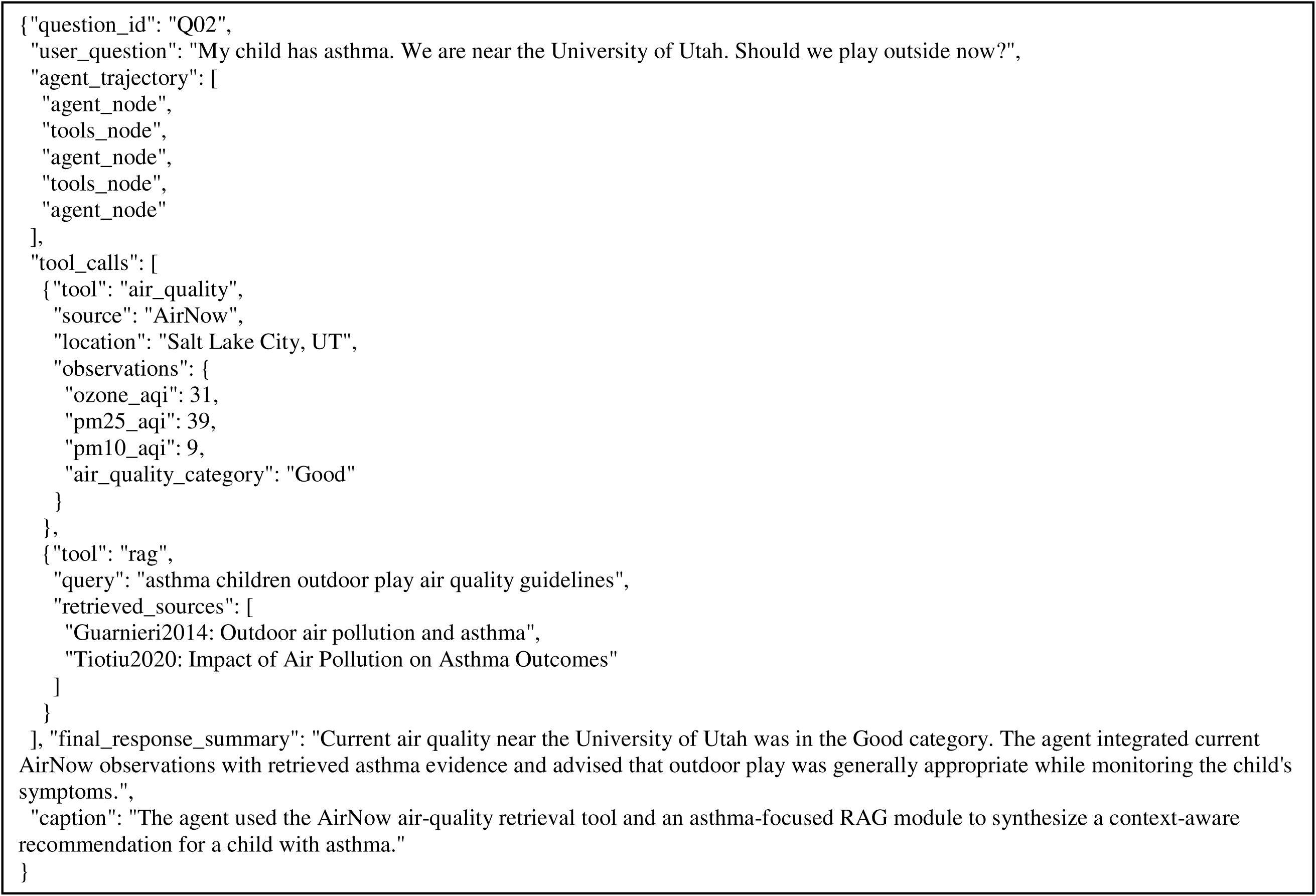
Agent execution trace for Q02: asthma-sensitive outdoor activity query near the University of Utah.

